# Contribution of Rare Large High-Penetrance CNVs to Pediatric and MODY Diabetes in Norway

**DOI:** 10.1101/2025.02.25.25322584

**Authors:** Haydee Artaza, Divya Sri Priyanka Tallapragada, Janne Molnes, Ksenia Lavrichenko, Anette S.B. Wolff, Ellen C. Røyrvik, Torild Skrivarhaug, Marc Vaudel, Eirik Bratland, Bente B. Johansson, Pål R. Njølstad, Stefan Johansson

## Abstract

Technological advancements have significantly improved our understanding of Copy Number Variants (CNVs) and their role in disease. However, detecting CNVs in clinical diagnostics remains challenging, and important pathogenic CNVs may go undetected.

This study systematically assessed the impact of rare, large, high-penetrance CNVs on pediatric diabetes and Maturity-onset Diabetes of the Young (MODY) in Norway. We analyzed data from the nationwide Norwegian Childhood Diabetes Registry (NCDR) covering 2002-2018 and the Norwegian MODY Registry (NMR) from 1997-2019. CNV detection was performed using the Illumina Infinium Global Screen Array-24 v2.0 on a total of 5,889 individuals and we compared the results to diagnostic records.

Our findings indicate that 0.63% of the patients in the Norwegian MODY Registry and 0.09% in the Norwegian Childhood Diabetes Registry are attributable to established pathogenic large copy number deletions detectable by array genotyping. Notably, six of the 14 pathogenic deletions identified (in the *HNF1B* [n=3], *HNF1A* [n=2], or *GATA4* genes [n=1]) had not been detected through standard diagnostic methods in the routine diagnostic screening. For these individuals, accurate molecular diagnoses have significant implications for personalized treatment and follow-up. We found no evidence suggesting a major role for additional rare CNVs beyond the already established pathogenic CNVs in MODY.

In conclusion, while pathogenic CNVs are rare, they remain relevant for patients of the Norwegian nationwide diabetes registries. Expanding screening for MODY variants, specifically 17q12-HNF1B and *HNF1A* deletions, to a larger portion of the pediatric diabetes population should be considered.

## Introduction

Diabetes is characterised by continued elevated blood glucose levels (1) due to pancreatic β-cell dysfunction causing insulin deficiency and/or resistance to insulin action (2,3). Accurate classification of diabetes is critical for guiding effective treatment strategies. However, distinguishing between diabetes subtypes can be challenging, particularly in younger adults, as many patients do not fit into established categories (2). In children and young adults without obesity, type 1 diabetes (T1D) and Maturity Onset Diabetes of the Young (MODY) are the two most common forms of diabetes, and these are often not correctly distinguished, having a possible impact on treatment choices.

T1D accounts for ∼5-10% of global diabetes cases (2,3) and is characterised by autoimmune-mediated beta-cell destruction, typically leading to absolute insulin deficiency (2,4). Autoantibodies, such as those targeting Glutamic Acid Decarboxylase (GADA), Insulinoma Antigen-2 (IA-2A), Insulin (IAA), and the Zinc Transporter 8 (ZnT8A) are often detectable before clinical diagnosis (4,5). MODY, the most common form of monogenic diabetes, results from mutations in one of at least ten known genes, accounting for ∼1-5% of diabetes cases (6). It typically presents in younger individuals (onset usually before 25-35 years of age) and is characterised by a progressive decline in insulin secretion, usually in the absence of autoantibodies and insulin resistance (7–9). The most common genetic forms of MODY are caused by mutations in the *GCK* (MODY2), *HNF1A* (MODY3), *HNF1B* (MODY5), and *HNF4A* (MODY1) genes (10). Diagnostic criteria for genetic screening of MODY include persistent hyperglycemia in early adulthood, clinical features incompatible with type 1 or type 2 diabetes mellitus, family history in at least one first-degree relative, evidence of residual pancreatic function, and the absence of beta-cell autoimmunity (7).

Genome-wide association studies (GWAS) have identified single nucleotide polymorphisms (SNPs) across ∼60 genomic regions that are associated with T1D risk in individuals with European ancestry (11–19). Collectively, these SNPs can explain around 80-85% of the T1D heritability (20). However, large genomic alterations such as copy number variations (CNVs), which range from a few kilobases (kb) to megabases (Mb), could account for the remaining 15–20% of unexplained heritability (20). Rare CNVs have been implicated in many human diseases, including neurodevelopmental and autoimmune disorders (21). Rare CNVs have also been associated with increased T1D susceptibility (22), and CNVs affecting MODY-related and other diabetes-associated genes like *GCK, HNF1B*, and *CEL* have been described as causal of MODY (23–25).

Differences in the genetic architecture between T1D and MODY could explain the distinct pathogenic mechanisms and differences in clinical presentation. While T1D is predominantly driven by autoimmune processes linked to the HLA region (26–28), MODY most commonly results from point mutations in genes critical for pancreatic function (29). Although large partial- or whole-gene deletions, including those in the *HNF1B, HNF1A* and *HNF4A* genes, have been observed in patients with suspected MODY where HNF1B deletions are most frequent (30,31), there is a lack of large empirical studies, that survey the prevalence of pathogenic CNVs in the general diabetes population without prior phenotypic selection. It is thus possible that MODY caused by CNVs are frequently underdiagnosed or misclassified as other forms of diabetes. Our study investigated the prevalence of rare CNVs and their potential diagnostic utility in a total of 5,889 patients from two nationwide Norwegian diabetes registries - the Norwegian Childhood Diabetes Registry (NCDR) covering nearly all children and adolescents with diabetes in Norway diagnosed from 2002 to 2018, and the Norwegian MODY Registry (NMR) in the time between 1997 to 2019.

## Materials and methods

### Study population

The Norwegian MODY Registry (NMR) was established in 1997 to serve as a national registry for patients with confirmed or clinical suspected monogenic diabetes, aimed at supporting both diagnostic and research efforts (32). The Norwegian Childhood Diabetes Registry (NCDR), operational since 2002, includes nearly all children and adolescents with type 1 diabetes in Norway (98% completeness) and collects comprehensive clinical data on children with diabetes, including family history, disease onset, treatment, and biochemical and genetic data (33). The study included all patients who had given consent to research for whom high-quality DNA was available and who had been recruited from 1997 to 2019 for the NMR and between 2002 and 2018 in NCDR in the A total of 1,584 individuals included in the NMR and 4,305 individuals included in the NCDR were genotyped using the Illumina Infinium Global Screen Array-24 v2.0.

### Genomic data quality control and CNV detection

A total of 741,187 markers were mapped to the GRCh37 assembly, and the intensity values of the autosomal SNP probes were extracted using GenomeStudio software version 2.0.4. CNVs were called using PennCNV (34) version 1.0.5. CNV calls overlapping at least 50% of genomic regions known to be problematic for CNV discovery –such as immunoglobulin, telomeric and centromeric regions– were excluded. CNVs fragmented into several adjacent small segments were merged into one single call if the gap length between them, divided by the total length of the resulting merged CNV call, was less than 50%. A second iteration of merging the CNV fragments with a stringent threshold of less than 40% was performed to identify robust CNV calls. Post-QC, we had 100,512 CNV calls, including 44,926 deletions and 55,586 duplications.

Low quality samples were excluded based on default quality metrics from PennCNV, including statistics for each sample: logR ratio standard deviation (LRR_SD <0.3), B allele frequency drift (BAF_drift <0.01) and waviness factor (|WF| <0.05). Additionally, we used the distribution of the total number of called CNVs (NumCNVs) per individual as a metric to identify samples of lesser quality. After carefully evaluating NumCNVs across all samples, we defined our exclusion threshold at NumCNVs >50 to ensure the high quality of the retained data and to avoid spurious calls. We excluded 844 samples based on these thresholds. Using PLINK (version 1.9), we calculated identity-by-descent (IBD) estimates to identify duplicates using a threshold of 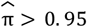. 95. We consolidated information from duplicated entries to retain a single entry. After excluding duplicate entries, we analysed data from a total of 5,045 individuals: 1,462 individuals from NMR and 3,583 individuals from NCDR, hereafter referred to as *NMR*_*ALL*_ and *NCDR*_*ALL*_, respectively.

### The unrelated-only, Northern European core subsets (*NCDR*_*CORE*_ and *NMR*_*CORE*_)

To allow for statistical comparison, we created a “core” subset from our overall dataset using the following filters, as depicted in Figure 1. First, to address genetic relatedness in the NMR dataset, particularly due to the referral of extended family members for screening to the NMR - we only included probands - typically the first family member referred for a molecular diagnosis. Then, we accounted for relatedness in the overall dataset by excluding samples with 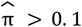. Also, nine individuals identified with trisomy 21 were excluded from the core. This yielded a subset of 685 individuals from NMR_ALL_ and 3,314 individuals from NCDR_ALL_. In these sub-cohorts, we restricted our analysis to unrelated individuals with a Northern European ancestry from both the registries and belonging to the following four subtypes: 1) individuals with a confirmed molecular diagnosis of MODY (**known MODY**); 2) individuals who meet the criteria for MODY—diagnosed at age ≤ 40 years (or assumed so if data is missing), with a normal BMI (≤ 30 kg/m^2^ or considered normal when unspecified), negative for autoantibodies (GADA, IA-2A, ZnT8A, or presumed negative if results are unavailable), and a family history of diabetes (diagnosed in at least two successive generations, or inferred as positive if information is lacking)—but do not have a confirmed molecular diagnosis (**likely MODY**); 3) individuals who tested positive for at least one autoantibody (autoantibodies against GADA, IA-2A and ZnT8A) at the time of referral (**classical T1D**); and 4) individuals who did not test positive for autoantibodies (Aab) at the time of diagnosis and did not fit the criteria for MODY (**Aab-negative T1D**). The resulting “core” dataset included a total of 3,798 individuals: 524 individuals from NMR and 3,274 individuals from the NCDR, hereafter referred to as *NMR*_*CORE*_ and *NCDR*_*CORE*,_ respectively.

**Figure 1.**
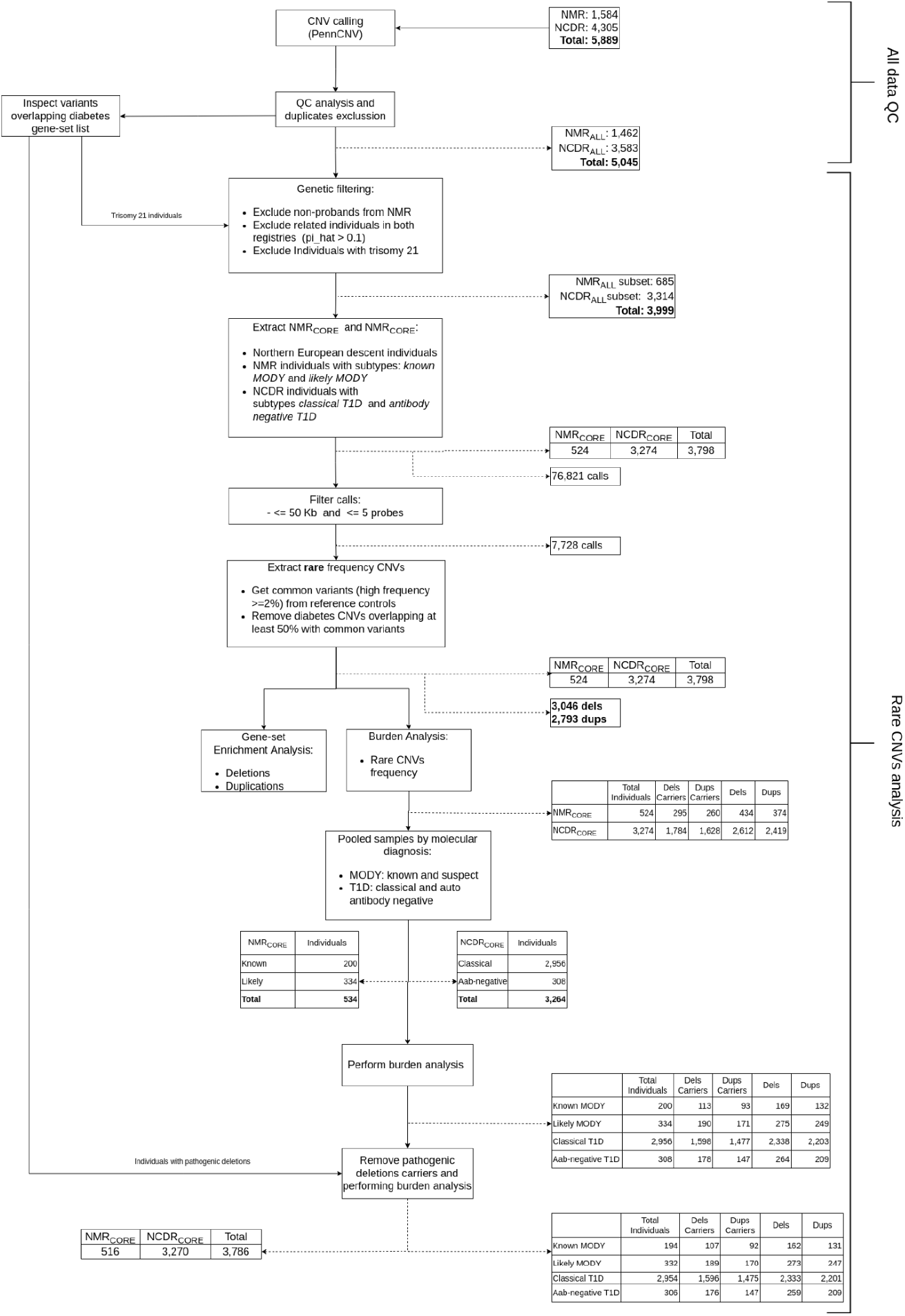
Flow chart of rare CNVs analysis in MODY and childhood-onset diabetes.

In the data analysis, we retained only large CNVs, spanning more than 50kb, covered by more than five probes. The final analysis included 7,728 CNVs in the “core” dataset with the following sample sizes: known MODY (N = 200), likely MODY (N = 334), classical T1D (N = 2,956) and Aab-negative T1D (N = 308).

### Rare CNVs

Common CNVs (carrier frequency >=2%) were identified using data from 200 healthy controls recruited through blood donor centers across Sweden and Norway as part of our previous study investigating the role of CNVs in Addison’s disease (35). CNVs from individuals in NMR_CORE_ and NCDR_CORE_ overlapping at least 50% with these common reference variants were classified as common variants and excluded from further analysis. Of 7,728 large CNVs, we retained 5,839 large, rare CNVs (carrier frequency < 2%), consisting of 3,046 rare deletions and 2,793 rare duplications, for subsequent analyses.

### Statistical analysis

We assessed the carrier frequencies of rare CNVs in our overall cohort comprising 5,045 individuals (*NMR*_*ALL*_: 1,462 and *NCDR*_*ALL*_: 3,583) and the core cohort comprising 3,798 individuals (*NMR*_*CORE*_: 524 and *NCDR*_*CORE*_: 3,274). To explore the distribution of CNVs by size, we categorised them into five interval ranges: 50–100 kb, 100–200 kb, 200–500 kb, 500–1000 kb, and >1,000 kb. We analysed cumulative frequencies for all CNVs and frequencies stratified by size intervals. Statistical differences between *NMR*_*ALL*_ and *NCDR*_*ALL*,_, and *NMR*_*CORE*_ and *NCDR*_*CORE*_ were evaluated using a two-proportion test, and the odds ratio estimation was performed using the *stats* and *fmsb* packages in R (version 3.4.4). We also performed burden analysis between the four subgroups: known MODY, likely MODY, classical T1D, and Aab-negative T1D. The burden analysis was subsequently repeated after excluding individuals with confirmed pathogenic rare CNVs to search for additional impact of other rare CNVs.

### Enrichment analysis

To test for potential enrichment of rare CNVs in diabetes-related genes, we performed enrichment analysis using a manually curated set of diabetes-related genes. We used the following gene sets derived from the Genomics England PanelApp (last accessed September 2023): Neonatal diabetes (V.4.0), Diabetes with additional phenotypes suggestive of a monogenic etiology (V.1.67), Familial diabetes (V.1.67), Insulin resistance (including lipodystrophy) (V.1.17), and Monogenic diabetes (V.2.2). We included an additional 16 genes implicated in early-onset diabetes and autoimmunity (36) to curate a gene list consisting of 98 unique genes (Supplementary Table A). Gene annotation for the entire genome was obtained from the UCSC Table Browser (build hg19/GRCh37) and downloaded via the PLINK resources page (32). We used Plink v.1.07 (37) to perform the analysis and used *--cnv-intersect* command to identify rare CNVs overlapping regions of the established Mendelian diabetes genes and used *--cnv-count, --cnv-subset*, and *--cnv-enrichment-test* commands to run gene-set enrichment analysis to identify enrichment of potentially pathogenic, rare CNVs in diabetes-related genes.

## Results

### Screening for established pathogenic CNVs increases diagnostic yield in the NCDR_ALL_ and NMR_ALL_ registries

We first screened for large chromosomal alterations known to be associated with diabetes among all available registry participants, categorised as NCDR_ALL_ (N = 3,583) and NMR_ALL_ (N = 1,462). In the NCDR_ALL_ group, nine individuals with trisomy 21 were identified, corresponding to a prevalence of 0.25% (9/3,583) in the Norwegian pediatric diabetes population (Supplementary Table 1). One individual (ID5) has a clinical picture more compatible with T2D, while the individual ID9 was first diagnosed with T1D and later was found to also carry a pathogenic GCK-MODY mutation. He presented with diabetes before the age of 10 with HbA1c of 15.7% (148 mmol/mol), BMI 12.5 kg/m^2^ and fasting serum C-peptide levels of 530 pmol/L, indicating a remission period with endogenous insulin production. He had detectable autoantibodies to GADA and IA-2A compatible with T1D and was hence treated with insulin.

**Table 1.** Pathogenic deletions in samples from the NCDR_ALL_ (n=1,462) and NMR_ALL_ (n=3,583) registries.

| Gene | CNV CHR:BP1-BP2 | Length (kb) | Reg. | Identified before* | Sub-cohort information | Subtype (prior to study) | Verified by |
| --- | --- | --- | --- | --- | --- | --- | --- |
| GATA4 | 8:8105359-11856903 | 3751.54 | NMR | No | Core** | Known MODY | ArrayCGH |
| HNF1A | 12:120974510-121712218 | 737.71 | NMR | No | Core** | Likely MODY | MLPA |
|  | 12:121426776-121435265 | 8.49 | NMR | No | Core: (deletion < 50 kb) | Likely MODY | MLPA |
|  | 12:121434074-121437802 | 3.73 | NMR | Yes | ALL-group-only: Non-NE ancestry sample (deletion < 50 kb) | Known MODY | MLPA |
| HNF1B | 17:34815551-36220373 | 1404.82 | NMR | Yes | Core** | Known MODY | MLPA |
|  | 17:34815551-36245768 | 1430.22 | NMR | Yes | Non-core: No proband | Known MODY | MLPA |
|  | 17:34815551-36245768 | 1430.22 | NMR | Yes | Core** | Known MODY | MLPA |
|  | 17:34815551-36249430 | 1433.88 | NMR | Yes | Core** | Known MODY | MLPA |
|  | 17:34815551-36249430 | 1433.88 | NMR | Yes | Core** | Known MODY | MLPA |
|  | 17:34815551-36249430 | 1433.88 | NMR | Yes | Core** | Known MODY | MLPA |
|  | 17:34528650-36220373 | 1691.72 | NCDR | No | Core** | IA-2A-positive T1D | MLPA |
|  | 17:34815551-36249430 | 1433.88 | NCDR | Yes | Core** | Aab-negative T1D | MLPA |
|  | 17:34815551-36249430 | 1433.88 | NCDR | No | Core** | Aab-negative T1D | MLPA |
|  | 17:34902695-36249430 | 1346.74 | NCDR | No | Core** | IA-2A-positive T1D | MLPA |

NMR: Norwegian MODY Registry; NCDR: Norwegian Childhood Diabetes Registry.
(\*) Was this deletion previously identified and documented in the Registry?
(\*\*) Carriers of pathogenic deletions included in $NMR_{CORE}$ and $NCDR_{CORE}$ analysis

Further screening for pathogenic deletions in established Mendelian diabetes genes revealed 14 CNVs with strong evidence of being pathogenic, of which six were not previously reported in any of the two registries, thus leading to new molecular diagnoses. All 14 variants were confirmed using orthogonal techniques (Table 1). The variants include one large 3.75 Mb deletion overlapping the *GATA4* locus in an individual from the NMR_ALL_ group. This individual presented with diabetes in his 10s with HbA1c of 10%, BMI 18.3 kg/m^2^, fasting serum C-peptide levels of 86 pmol/L and no detectable levels of T1D autoantibodies (Supplementary Table 2). Exocrine pancreatic function was unknown. Other notable clinical features include congenital hernia and heart defects consistent with *GATA4* haploinsufficiency (38). He was treated with insulin. Three deletions overlapping *HNF1A* (one whole gene deletion and two partial gene deletions) were identified in NMR_ALL_, of which only one was previously known to the patient. All three individuals with *HNF1A* deletions had autoantibody-negative diabetes and a family history of diabetes over three generations. Moreover, ten individuals, including four males and six females, with *HNF1B* deletions were identified. Deletions in six of these individuals were previously reported in NMR_ALL_, where five of them showed extra-pancreatic features, and none were positive for T1D-autoantibodies (Supplementary Table 2). The remaining four individuals with *HNF1B* deletions were first reported in NCDR_ALL_ of which only one had been detected independently of this study (Table 1). Two of these newly detected deletions were identified in individuals positive for IA-2A autoantibodies at diagnosis and treated with insulin, and the third deletion was identified in an individual without detectable T1D-autoantibodies treated with insulin and presenting with unspecified liver disease and allergy (Supplementary Table 2). The presence of these pathogenic deletions confirms the molecular diagnosis of diabetes in these individuals as HNF1B-MODY.

**Table 2.** Distribution of rare CNVs in *NMR*_*CORE*_ and *NCDR*_*CORE*_.

| Overlapped Region |  | CNVs<br>NMR<br>[524] | CNVs<br>NCDR<br>[3274] | Frequency |  | Association |  |
| --- | --- | --- | --- | --- | --- | --- | --- |
|  |  |  |  | NMR | NCDR | OR (95%CI) | P |
| (a) Genic | DELs | 434 | 2612 | 0.83 | 0.80 | 1.22 (0.96, 1.56) | 0.10 |
|  | DUPs | 374 | 2419 | 0.71 | 0.74 | 0.88 (0.72, 1.08) | 0.23 |
| (b) Diabetes<br>gene-set | DELs | 7 | 9 | 0.01 | 0.003 | 4.91 (1.82,13.25) | 4.99E-04 |
|  | DUPs | 2 | 10 | 0.004 | 0.003 | 1.25 (0.27-5.72) | 0.77 |

### CNV analyses in non-related individuals of Northern European descent

For subsequent analyses, we excluded individuals with trisomy 21 and, to allow for statistical comparison, we restricted the investigation to unrelated individuals of Northern European ancestry, forming two subsets *NCDR*_*CORE*_ (N = 3,274) and *NMR*_*CORE*_ (N= 524), respectively, as outlined in Figure 1. We analysed large, rare CNVs, defined as > 50 kb in size, supported by > five probes with a carrier frequency of <2% in controls. This yielded a total of 3,046 rare deletions and 2,793 rare duplications. A global burden analysis showed no nominally significant differences in the overall distribution of rare deletions or duplications between individuals in the *NMR*_*CORE*_ and *NCDR*_*CORE*_ groups (Table 2-a). However, when stratified by CNV size, a few nominally significant findings emerged: deletions in the 50–100 kb range were more frequent in the *NMR*_*CORE*_ group (OR = 1.21, 95% CI: 1.00–1.45, *p* = 0.05; Table 3), whereas duplications in the same size range were more common in the *NCDR*_*CORE*_ group (OR = 0.75, 95% CI: 0.61–0.93, *p* = 0.01; Table 3). Additionally, deletions exceeding 1,000 kb appeared more frequent in the *NMR*_*CORE*_ group (OR = 2.27, 95% CI: 1.05–4.89, *p* = 0.03; Table 3). However, none of these findings remained significant after correction for the number of tests performed in this analysis (N=12).

**Table 3.** Distribution of rare CNVs within size intervals in *NMR*_*CORE*_ and *NCDR*_*CORE*_.

| CNVs length | | CNVs<br>$NMR_{CORE}$<br>[524] | CNVs<br>$NCDR_{CORE}$<br>[3274] | Frequency | | Association | |
| --- | --- | --- | --- | --- | --- | --- | --- |
| | | | | $NMR_{CORE}$ | $NCDR_{CORE}$ | OR (95%CI) | P |
| DELs | 50kb_100kb | 246 | 1386 | 0.47 | 0.42 | 1.21 (1.00, 1.45) | 0.05 |
|  | 100kb_200kb | 118 | 846 | 0.23 | 0.26 | 0.83 (0.67, 1.04) | 0.10 |
|  | 200kb_500kb | 53 | 300 | 0.10 | 0.09 | 1.12 (0.82, 1.52) | 0.49 |
|  | 500kb_1000kb | 8 | 55 | 0.02 | 0.02 | 0.91 (0.43, 1.92) | 0.80 |
|  | 1000kb_> | 9 | 25 | 0.02 | 0.01 | 2.27 (1.05, 4.89) | 0.03 |
| DUPs | 50kb_100kb | 135 | 1032 | 0.26 | 0.32 | 0.75 (0.61, 0.93) | 0.01 |
|  | 100kb_200kb | 113 | 693 | 0.22 | 0.21 | 1.02 (0.82, 1.28) | 0.84 |
|  | 200kb_500kb | 95 | 527 | 0.18 | 0.16 | 1.15 (0.91, 1.47) | 0.24 |
|  | 500kb_1000kb | 23 | 126 | 0.04 | 0.04 | 1.15 (0.74, 1.81) | 0.55 |
|  | 1000kb_> | 8 | 41 | 0.02 | 0.01 | 1.22 (0.57, 2.62) | 0.61 |

Next, we restricted the analyses to a curated set of 98 diabetes-related genes (Supplementary Table A). Here, we found a higher frequency of deletions overlapping diabetes-related genes in individuals from the *NMR*_*CORE*_ group compared to the *NCDR*_*CORE*_ group (OR = 4.91, 95% CI: 1.82–13.25, p = 4.99E-04; Table 2-b). These CNVs include the *GATA4* deletion, the large *HNF1A* gene deletion, and nine *HNF1B* deletions described above (marked with ** in Table 1). To investigate if there is any evidence of additional CNV burden beyond the clinically significant deletions, we excluded the 11 individuals harboring these deletions in MODY-associated genes (denoted with ** in Table 1) and repeated our analyses. After this exclusion, neither the global burden analysis nor the diabetes gene set analysis revealed any notable differences in the distribution of CNVs between the groups (Supplementary Table 3). Some tendencies for the smaller CNVs (50–100 kb) remained evident (Supplementary Table 4). Combined, these findings do not support a broader enrichment of not already established diabetes-related deletions in the *NMR*_*CORE*_ group compared to children recruited from the overall pediatric diabetes population (*NCDR*_*CORE*_).

### CNV analyses in clinically defined subtypes of the NCDR_CORE_ and NMR_CORE_ registries

It can be speculated that rare CNVs might contribute differently to different subtypes within the two registries. Although power is low when sub-stratifying the cohorts further, we performed an explorative overview of the burden of rare CNVs across four subtypes of individuals within the *NCDR*_*CORE*_ and *NMR*_*CORE*_ cohorts: Individuals with a confirmed molecular MODY diagnosis (“known MODY”: N = 194), and individuals without a known molecular cause but fulfilling clinical MODY criteria (“likely MODY”: N = 332), “classical T1D” with presence of auto-antibodies (N = 2,954), and Aab-negative T1D (N = 306). Results are shown in Tables 4 and 5 and Supplementary Table 5. Some nominal associations were seen, but none of the findings remained significant after Bonferroni correction for multiple tests.

**Table 4.** Distribution of rare CNVs in individuals within the subtypes of *classical* T1D and *autoantibody-negative* T1D.

|  | CNVs<br>Classical T1D<br>[2954] | CNVs<br>Aab-negative T1D<br>[306] | Frequency |  | Association |  |
| --- | --- | --- | --- | --- | --- | --- |
|  |  |  | Classical<br>T1D | Aab-negative<br>T1D | OR (95%CI) | *P* |
| DELS | 2333 | 259 | 0.79 | 0.85 | 0.68 (0.49-0.94) | 0.02 |
| DUPs | 2201 | 209 | 0.75 | 0.68 | 1.36 (1.05-1.75) | 0.02 |

**Table 5.** Distribution of rare CNVs for individuals within the subtypes of *known MODY* and *classical T1D*.

|  | CNVs<br>Known MODY<br>[194] | CNVs<br>Classical T1D<br>[2954] | Frequency |  | Association |  |
| --- | --- | --- | --- | --- | --- | --- |
|  |  |  | Known<br>MODY | Classical<br>T1D | OR (95%CI) | *P* |
| DELS | 162 | 2333 | 0.84 | 0.79 | 1.35 (0.91-1.99) | 0.13 |
| DUPs | 131 | 2201 | 0.68 | 0.75 | 0.71 (0.52-0.97) | 0.03 |

We have also listed the distribution of rare deletions (after excluding the known pathogenic deletions) by the most common genetic subtypes of MODY: MODY2 (*GCK*), MODY1 (*HNF4A*), and MODY5 (*HNF1B*), which represent the most prevalent monogenic etiologies (39) (Supplementary Table 6). As expected, no clear differences between subtypes were seen.

## Discussion

Traditionally, genetic screening has been reserved for pediatric and young adult patients presenting with atypical diabetes or when there is high suspicion of Mendelian diabetes. Sophisticated clinical algorithms, such as the Exeter MODY calculator (40) and expert recommendations (27,41), are constantly updated to aid clinical decision-making. However, these algorithms typically incorporate the segregation of diabetes across multiple generations and clinical characteristics, which may overlook individuals with *de novo* pathogenic variants and atypical clinical presentations. These two features are particularly relevant to and often characterize large, rare pathogenic CNVs. Moreover, conventional screening approaches like Sanger sequencing and whole-exome sequencing are not optimised for detecting CNVs, limiting their ability to systematically identify potentially pathogenic structural variants (42,43). Consequently, most clinical laboratories have largely restricted CNV analyses to selected patient subsets, thus limiting the detection of pathogenic CNVs.

Here, we show that 0.63% of cases in the Norwegian nationwide MODY registry (NMR) (n=10/1,584 individuals) and 0.09% of cases in the nationwide childhood diabetes registry (NCDR) (n=4/4,305) can be ascribed to established pathogenic large deletions detectable by standard array genotyping. Notably, six of the 14 (43%) pathogenic deletions had not been detected by the standard of care in the Norwegian health system, thus including CNV analysis in the diagnostic repertoire enables additional molecular diagnoses with potential implications for personalized treatment and follow-up.

In our systematic analysis of the broader NMR_ALL_ and NCDR_ALL_ cohorts, we identified nine cases of trisomy 21, corresponding to a prevalence of 0.25% in the pediatric diabetes population. These findings align with previous evidence indicating that individuals with trisomy 21 have an increased risk for diabetes (44–46). We also identified a 3.75-Mb deletion overlapping the *GATA4* gene in an individual with congenital hernia and heart defects, features typically associated with *GATA4* haploinsufficiency (38,47,48).

Our study identified four of 3,583 children with diabetes (∼1 in 900) in the NCDR_ALL_ cohort with large deletions at the *HNF1B* locus. As three of these deletions had not been discovered prior to this study, a molecular diagnosis of HNF1B-MODY could now be assigned for these three individuals. Mutations in the *HNF1B* gene and whole-gene deletions as a part of a larger 17q12 deletion syndrome are well-established causes of HNF1B-MODY and are usually associated with extra-pancreatic features such as renal cysts (49–51). Although HNF1B-MODY typically follows an autosomal dominant inheritance pattern, up to 50% of deletions in *HNF1B* are reported to occur *de novo* (52,53). Our findings align with these reports as there were no indications of parental diabetes for these individuals in the registry data. We cannot rule out under-reporting of data in the registry, but it underscores the importance of molecular testing for *HNF1B* mutations in patients presenting with diabetes and extrapancreatic manifestations, even in the absence of family history. These results demonstrate the clinical utility of CNVs in providing a molecular diagnosis of diabetes, particularly in *de novo* cases, which could inform personalized diabetes management and improve patient outcomes.

We did not find evidence for a contribution of large rare CNVs beyond those affecting *HNF1B* and *HNF1A*, neither in suspected MODY patients without a confirmed molecular diagnosis nor in autoantibody-positive and -negative childhood diabetes. However, the relatively small size of key subgroups, particularly in the NMR, limited our statistical power to detect evidence of novel pathogenic CNVs. Combining data from other MODY registries could help mitigate these constraints by increasing cohort size, enhancing statistical power, and enabling more comprehensive analyses of CNV contributions across diverse populations.

Another limitation is that smaller CNVs affecting single exons or smaller genomic regions will have been missed due to the limited resolution of the arrays employed in this study. We used a standard biobank scale GWAS array with limited ability to identify smaller deletions or duplications that might disrupt single exons or regulatory elements critical to gene function. Established technologies like whole-genome sequencing (41,54–56) or targeted methods like multiplex ligation-dependent probe amplification (MLPA) can enhance detection sensitivity for specific genes of interest by accurately sizing and quantifying smaller CNVs. In this regard, our preliminary analysis, unconstrained by CNV size, revealed two small deletions in the *HNF1A* gene (8.5 kb and 3.7 kb) which were subsequently confirmed via MLPA. Incorporating more high-resolution methods in future studies could improve diagnostic yield and refine our understanding of how structural genetic variation contributes to diabetes susceptibility and its clinical subtypes.

The discovery of CNVs in *HNF1A* or *HNF1B* in apparent T1D that we have described here has a major impact on clinical treatment. In HNF1A-MODY and HNF1B-MODY, patients can often be transferred from insulin to oral sulfonylurea or a combination of sulfonylurea and either a GLP1 or an SGLT2 analogue. Furthermore, a diagnosis of HNF1A-MODY or HNF1B-MODY has a major impact on genetic counselling and prognosis. Hence, this shows that it is warranted to screen all antibody-negative patients with apparent T1D for CNVs in *HNF1A* and *HNF1B*. It should also be noted that two of the *HNF1B*-deletion carriers were also positive for IA-2A-autoantibodies, hence it could be argued that extended screening may also include auto-antibody negative diabetes with detectable C-peptide levels.

Overall, our study contributes to the growing understanding of the complex genetic architecture of diabetes while exploring the utility of CNV analysis in precision diagnostics of diabetes and highlights the potential to refine diabetes classification and management, paving the way for personalized medicine in diabetes care.

## Data Availability

The datasets presented in this article are not readily available because of restrictions related to ethical approvals regarding sharing of human genomic data. Requests to access the datasets should be directed to PRN. Access to the dataset requires an ethical approval from the requesting party.

## Ethics Statement

The study was approved by the Regional Committee for Medical Research Ethics, (REK-Vest no. 2009/2079, 2009/2080, 2013/1504 and 2017/624).

## Funding and Acknowledgments

We are thankful to all the individuals with diabetes and their families. We thank the Norwegian Study Group for Childhood and Adolescent Diabetes for providing data to the NCDR.

This work was supported by grants (to S.J) Helse Vest’s Open Research Grant (grants #912250) and F-12144), the Novo Nordisk Foundation (grantNNF20OC0063872), the Research Council of Norway (grant #315599), and (to A.S.B.W) the Novo Nordisk Fonden NNF240C0088909 and Western Norway Health Authorities

## Author contributions

H.A, D.S.P.T and S.J. wrote the manuscript. S.J. designed the study. H.A and D.S.P.T analysed the data. J.M., A.S.B.W., T.S., B.B.J and P.R.N contributed to sample acquisition and clinical information. K.L., E.C.R., M.V, E.B., P.R.N and S.J. provided statistical and technical advice. All authors participated in preparing the manuscript by reading and commenting on drafts before submission. P.R.N. and S.J. acquired the funding.

## References

1. Banday MZ, Sameer AS, Nissar S. Pathophysiology of diabetes: An overview. Avicenna J Med (2020) 10:174–188. doi: 10.4103/ajm.ajm_53_20

2. American Diabetes Association. Diagnosis and Classification of Diabetes Mellitus. Diabetes Care (2013) 37:S81–S90. doi: 10.2337/dc14-S081

3. Skyler JS, Bakris GL, Bonifacio E, Darsow T, Eckel RH, Groop L, Groop P-H, Handelsman Y, Insel RA, Mathieu C, et al. Differentiation of Diabetes by Pathophysiology, Natural History, and Prognosis. Diabetes (2017) 66:241–255. doi: 10.2337/db16-0806

4. Daneman D. Type 1 diabetes. The Lancet (2006) 367:847–858. doi: 10.1016/S0140-6736(06)68341-4

5. Wenzlau JM, Frisch LM, Gardner TJ, Sarkar S, Hutton JC, Davidson HW. Novel antigens in type 1 diabetes: The importance of ZnT8. Curr Diab Rep (2009) 9:105–112. doi: 10.1007/s11892-009-0019-4

6. Zhang H, Colclough K, Gloyn AL, Pollin TI. Monogenic diabetes: a gateway to precision medicine in diabetes. J Clin Invest 131:e142244. doi: 10.1172/JCI142244

7. Nkonge KM, Nkonge DK, Nkonge TN. The epidemiology, molecular pathogenesis, diagnosis, and treatment of maturity-onset diabetes of the young (MODY). Clin Diabetes Endocrinol (2020) 6:20. doi: 10.1186/s40842-020-00112-5

8. Molven A, NjøLstad PR. Role of molecular genetics in transforming diagnosis of diabetes mellitus. Expert Rev Mol Diagn (2011) 11:313–20. doi: 10.1586/erm.10.123

9. De Franco E. From Biology to Genes and Back Again: Gene Discovery for Monogenic Forms of Beta-Cell Dysfunction in Diabetes. J Mol Biol (2020) 432:1535–1550. doi: 10.1016/j.jmb.2019.08.016

10. Johansson S, Irgens H, Chudasama KK, Molnes J, Aerts J, Roque FS, Jonassen I, Levy S, Lima K, Knappskog PM, et al. Exome Sequencing and Genetic Testing for MODY. PLoS ONE (2012) 7:e38050. doi: 10.1371/journal.pone.0038050

11. Todd JA, Walker NM, Cooper JD, Smyth DJ, Downes K, Plagnol V, Bailey R, Nejentsev S, Field SF, Payne F, et al. Robust associations of four new chromosome regions from genome-wide analyses of type 1 diabetes. Nat Genet (2007) 39:857–864. doi: 10.1038/ng2068

12. Cooper JD, Smyth DJ, Smiles AM, Plagnol V, Walker NM, Allen J, Downes K, Barrett JC, Healy B, Mychaleckyj JC, et al. Meta-analysis of genome-wide association study data identifies additional type 1 diabetes loci. Nat Genet (2008) 40:1399–1401. doi: 10.1038/ng.249

13. Hakonarson H, Qu H-Q, Bradfield JP, Marchand L, Kim CE, Glessner JT, Grabs R, Casalunovo T, Taback SP, Frackelton EC, et al. A novel susceptibility locus for type 1 diabetes on Chr12q13 identified by a genome-wide association study. Diabetes (2008) 57:1143–1146. doi: 10.2337/db07-1305

14. Barrett JC, Clayton D, Concannon P, Akolkar B, Cooper JD, Erlich HA, Julier C, Morahan G, Nerup J, Nierras C, et al. Genome-wide association study and meta-analysis finds over 40 loci affect risk of type 1 diabetes. Nat Genet (2009) 41:703–707. doi: 10.1038/ng.381

15. Grant SFA, Qu H-Q, Bradfield JP, Marchand L, Kim CE, Glessner JT, Grabs R, Taback SP, Frackelton EC, Eckert AW, et al. Follow-Up Analysis of Genome-Wide Association Data Identifies Novel Loci for Type 1 Diabetes. Diabetes (2009) 58:290–295. doi: 10.2337/db08-1022

16. Bradfield JP, Qu H-Q, Wang K, Zhang H, Sleiman PM, Kim CE, Mentch FD, Qiu H, Glessner JT, Thomas KA, et al. A Genome-Wide Meta-Analysis of Six Type 1 Diabetes Cohorts Identifies Multiple Associated Loci. PLOS Genet (2011) 7:e1002293. doi: 10.1371/journal.pgen.1002293

17. Huang J, Ellinghaus D, Franke A, Howie B, Li Y. 1000 Genomes-based imputation identifies novel and refined associations for the Wellcome Trust Case Control Consortium phase 1 Data. Eur J Hum Genet (2012) 20:801–805. doi: 10.1038/ejhg.2012.3

18. Zhu M, Xu K, Chen Y, Gu Y, Zhang M, Luo F, Liu Y, Gu W, Hu J, Xu H, et al. Identification of Novel T1D Risk Loci and Their Association With Age and Islet Function at Diagnosis in Autoantibody-Positive T1D Individuals: Based on a Two-Stage Genome-Wide Association Study. Diabetes Care (2019) 42:1414–1421. doi: 10.2337/dc18-2023

19. Onengut-Gumuscu S, Chen W-M, Burren O, Cooper NJ, Quinlan AR, Mychaleckyj JC, Farber E, Bonnie JK, Szpak M, Schofield E, et al. Fine mapping of type 1 diabetes susceptibility loci and evidence for colocalization of causal variants with lymphoid gene enhancers. Nat Genet (2015) 47:381–386. doi: 10.1038/ng.3245

20. Pang H, Lin J, Luo S, Huang G, Li X, Xie Z, Zhou Z. The missing heritability in type 1 diabetes. Diabetes Obes Metab (2022) 24:1901–1911. doi: 10.1111/dom.14777

21. Auwerx C, Jõeloo M, Sadler MC, Tesio N, Ojavee S, Clark CJ, Mägi R, Reymond A, Kutalik Z. Rare copy-number variants as modulators of common disease susceptibility. Genome Med (2024) 16:5. doi: 10.1186/s13073-023-01265-5

22. Cooper NJ, Shtir CJ, Smyth DJ, Guo H, Swafford AD, Zanda M, Hurles ME, Walker NM, Plagnol V, Cooper JD, et al. Detection and correction of artefacts in estimation of rare copy number variants and analysis of rare deletions in type 1 diabetes. Hum Mol Genet (2015) 24:1774–1790. doi: 10.1093/hmg/ddu581

23. Yu R, Zhang H, Xiao X. Partial GCK gene deletion mutations causing maturity-onset diabetes of the young. Acta Diabetol (2024) 61:107–115. doi: 10.1007/s00592-023-02173-1

24. Berberich AJ, Wang J, Cao H, McIntyre AD, Spaic T, Miller DB, Stock S, Huot C, Stein R, Knoll J, et al. Simplifying Detection of Copy-Number Variations in Maturity-Onset Diabetes of the Young. Can J Diabetes (2021) 45:71–77. doi: 10.1016/j.jcjd.2020.06.001

25. Torsvik J, Johansson S, Johansen A, Ek J, Minton J, Ræder H, Ellard S, Hattersley A, Pedersen O, Hansen T, et al. Mutations in the VNTR of the carboxyl-ester lipase gene (CEL) are a rare cause of monogenic diabetes. Hum Genet (2010) 127:55–64. doi: 10.1007/s00439-009-0740-8

26. Davies JL, Kawaguchi Y, Bennett ST, Copeman JB, Cordell HJ, Pritchard LE, Reed PW, Gough SCL, Jenkins SC, Palmer SM, et al. A genome-wide search for human type 1 diabetes susceptibility genes. Nature (1994) 371:130–136. doi: 10.1038/371130a0

27. Concannon P, Gogolin-Ewens KJ, Hinds DA, Wapelhorst B, Morrison VA, Stirling B, Mitra M, Farmer J, Williams SR, Cox NJ, et al. A second-generation screen of the human genome for susceptibility to insulin-dependent diabetes mellitus. Nat Genet (1998) 19:292–296. doi: 10.1038/985

28. Concannon P, Erlich HA, Julier C, Morahan G, Nerup J, Pociot F, Todd JA, Rich SS, the Type 1 Diabetes Genetics Consortium. Type 1 Diabetes: Evidence for Susceptibility Loci from Four Genome-Wide Linkage Scans in 1,435 Multiplex Families. Diabetes (2005) 54:2995–3001. doi: 10.2337/diabetes.54.10.2995

29. Sanyoura M, Philipson LH, Naylor R. Monogenic Diabetes in Children and Adolescents: Recognition and Treatment Options. Curr Diab Rep (2018) 18:58. doi: 10.1007/s11892-018-1024-2

30. Clissold RL, Ashfield B, Burrage J, Hannon E, Bingham C, Mill J, Hattersley A, Dempster EL. Genome-wide methylomic analysis in individuals with HNF1B intragenic mutation and 17q12 microdeletion. Clin Epigenetics (2018) 10:97. doi: 10.1186/s13148-018-0530-z

31. Colclough K, Ellard S, Hattersley A, Patel K. Syndromic monogenic diabetes genes should be tested in patients with a clinical suspicion of MODY. Diabetes (2022) 71:530–537. doi: 10.2337/db21-0517

32. Søvika O, Irgens HU, Molnes J, Sagena JV, Bjørkhaug L, Ræder H, Molveng A, Njølstad PR. Monogenic diabetes mellitus in Norway. Nor Epidemiol (2013) 23: doi: 10.5324/nje.v23i1.1603

33. Skrivarhaug T. Norwegian Childhood Diabetes Registry: Childhood onset diabetes in Norway 1973-2012. Nor Epidemiol (2013) 23: doi: 10.5324/nje.v23i1.1598

34. Wang K, Li M, Hadley D, Liu R, Glessner J, Grant SFA, Hakonarson H, Bucan M. PennCNV: An integrated hidden Markov model designed for high-resolution copy number variation detection in whole-genome SNP genotyping data. Genome Res (2007) 17:1665–1674. doi: 10.1101/gr.6861907

35. Artaza H, Eriksson D, Lavrichenko K, Aranda-Guillén M, Bratland E, Vaudel M, Knappskog P, Husebye ES, Bensing S, Wolff ASB, et al. Rare copy number variation in autoimmune Addison’s disease. Front Immunol (2024) 15:1374499. doi: 10.3389/fimmu.2024.1374499

36. Johnson MB, Hattersley AT, Flanagan SE. Monogenic autoimmune diseases of the endocrine system. Lancet Diabetes Endocrinol (2016) 4:862–872. doi: 10.1016/S2213-8587(16)30095-X

37. Raychaudhuri S, Korn JM, McCarroll SA, Consortium TIS, Altshuler D, Sklar P, Purcell S, Daly MJ. Accurately Assessing the Risk of Schizophrenia Conferred by Rare Copy-Number Variation Affecting Genes with Brain Function. PLOS Genet (2010) 6:e1001097. doi: 10.1371/journal.pgen.1001097

38. Shaw-Smith C, De Franco E, Lango Allen H, Batlle M, Flanagan SE, Borowiec M, Taplin CE, van Alfen-van der Velden J, Cruz-Rojo J, Perez de Nanclares G, et al. GATA4 mutations are a cause of neonatal and childhood-onset diabetes. Diabetes (2014) 63:2888–2894. doi: 10.2337/db14-0061

39. Kavvoura FK, Owen KR. Maturity onset diabetes of the young: clinical characteristics, diagnosis and management. Pediatr Endocrinol Rev PER (2012) 10:234–242.

40. Shields BM, McDonald TJ, Ellard S, Campbell MJ, Hyde C, Hattersley AT. The development and validation of a clinical prediction model to determine the probability of MODY in patients with young-onset diabetes. Diabetologia (2012) 55:1265–1272. doi: 10.1007/s00125-011-2418-8

41. Murphy R, Colclough K, Pollin TI, Ikle JM, Svalastoga P, Maloney KA, Saint-Martin C, Molnes J, Misra S, Aukrust I, et al. The use of precision diagnostics for monogenic diabetes: a systematic review and expert opinion. Commun Med (2023) 3:1–24. doi: 10.1038/s43856-023-00369-8

42. Zare F, Dow M, Monteleone N, Hosny A, Nabavi S. An evaluation of copy number variation detection tools for cancer using whole exome sequencing data. BMC Bioinformatics (2017) 18:286. doi: 10.1186/s12859-017-1705-x

43. Pös O, Radvanszky J, Styk J, Pös Z, Buglyó G, Kajsik M, Budis J, Nagy B, Szemes T. Copy Number Variation: Methods and Clinical Applications. Appl Sci (2021) 11:819. doi: 10.3390/app11020819

44. Hom B, Boyd NK, Vogel BN, Nishimori N, Khoshnood MM, Jafarpour S, Nagesh D, Santoro JD. Down Syndrome and Autoimmune Disease. Clin Rev Allergy Immunol (2024) 66:261–273. doi: 10.1007/s12016-024-08996-2

45. Aslam AA, Baksh RA, Pape SE, Strydom A, Gulliford MC, Chan LF, GO-DS21 Consortium. Diabetes and Obesity in Down Syndrome Across the Lifespan: A Retrospective Cohort Study Using U.K. Electronic Health Records. Diabetes Care (2022) 45:2892–2899. doi: 10.2337/dc22-0482

46. Aitken RJ, Mehers KL, Williams AJ, Brown J, Bingley PJ, Holl RW, Rohrer TR, Schober E, Abdul-Rasoul MM, Shield JPH, et al. Early-onset, coexisting autoimmunity and decreased HLA-mediated susceptibility are the characteristics of diabetes in Down syndrome. Diabetes Care (2013) 36:1181–1185. doi: 10.2337/dc12-1712

47. Ding L, Cai M, Chen L, Yan H, Lu S, Pang S, Yan B. Identification and functional study of GATA4 gene regulatory variants in type 2 diabetes mellitus. BMC Endocr Disord (2021) 21:73. doi: 10.1186/s12902-021-00739-0

48. Sartori DJ, Wilbur CJ, Long SY, Rankin MM, Li C, Bradfield JP, Hakonarson H, Grant SFA, Pu WT, Kushner JA. GATA Factors Promote ER Integrity and β-Cell Survival and Contribute to Type 1 Diabetes Risk. Mol Endocrinol (2014) 28:28–39. doi: 10.1210/me.2013-1265

49. Laffargue F, Bourthoumieu S, Llanas B, Baudouin V, Lahoche A, Morin D, Bessenay L, De Parscau L, Cloarec S, Delrue M-A, et al. Towards a new point of view on the phenotype of patients with a 17q12 microdeletion syndrome. Arch Dis Child (2015) 100:259–264. doi: 10.1136/archdischild-2014-306810

50. Roehlen N, Hilger H, Stock F, Gläser B, Guhl J, Schmitt-Graeff A, Seufert J, Laubner K. 17q12 Deletion Syndrome as a Rare Cause for Diabetes Mellitus Type MODY5. J Clin Endocrinol Metab (2018) 103:3601–3610. doi: 10.1210/jc.2018-00955

51. Clissold RL, Hamilton AJ, Hattersley AT, Ellard S, Bingham C. HNF1B-associated renal and extra-renal disease-an expanding clinical spectrum. Nat Rev Nephrol (2015) 11:102–112. doi: 10.1038/nrneph.2014.232

52. Edghill EL, Bingham C, Ellard S, Hattersley AT. Mutations in hepatocyte nuclear factor-1β and their related phenotypes. J Med Genet (2006) 43:84–90. doi: 10.1136/jmg.2005.032854

53. Hwang D-Y, Dworschak GC, Kohl S, Saisawat P, Vivante A, Hilger AC, Reutter HM, Soliman NA, Bogdanovic R, Kehinde EO, et al. Mutations in 12 known dominant disease-causing genes clarify many congenital anomalies of the kidney and urinary tract. Kidney Int (2014) 85:1429–1433. doi: 10.1038/ki.2013.508

54. Johansson BB, Irgens HU, Molnes J, Sztromwasser P, Aukrust I, Juliusson PB, Søvik O, Levy S, Skrivarhaug T, Joner G, et al. Targeted next-generation sequencing reveals MODY in up to 6.5% of antibody-negative diabetes cases listed in the Norwegian Childhood Diabetes Registry. Diabetologia (2017) 60:625–635. doi: 10.1007/s00125-016-4167-1

55. Maltoni G, Franceschi R, Di Natale V, Al-Qaisi R, Greco V, Bertorelli R, De Sanctis V, Quattrone A, Mantovani V, Cauvin V, et al. Next Generation Sequencing Analysis of MODY-X Patients: A Case Report Series. J Pers Med (2022) 12:1613. doi: 10.3390/jpm12101613

56. Al-Kandari H, Al-Abdulrazzaq D, Davidsson L, Nizam R, Jacob S, Melhem M, John SE, Al-Mulla F. Identification of Maturity-Onset-Diabetes of the Young (MODY) mutations in a country where diabetes is endemic. Sci Rep (2021) 11:16060. doi: 10.1038/s41598-021-95552-z

