## Supplementary Tables for "Contribution of Rare Large High-Penetrance CNVs to Pediatric and MODY Diabetes in Norway"

Table A. List of panels and genes included in gene-set enrichment analysis

diabetes_genes

Supplementary Table 1. Clinical Presentation of Individuals with Trisomy 21 in $NCDR_{ALL}$

| ID | Registry | Sub type | Sex | Age range* (years) | BMI (kg/m2) | Aa** | HbA1c (%) | C-peptide (pmol/l) | Treatment at discharge | Family History of diabetes |
|---|---|---|---|---|---|---|---|---|---|---|
| 1 | NCDR | Classical T1D | m | [0-4] | 15.11 | Positive for GADA, IA-2A | 10.5 | N/A | Insulin | N/A |
| 2 | NCDR | Classical T1D | m | [5-9] | 15.02 | Positive for GADA, IA-2A | 10.5 | N/A | Insulin | Paternal grandfather |
| 3 | NCDR | Classical T1D | m | [5-9] | 14.40 | Positive for GADA | 12.7 | 213 | Insulin | N/A |
| 4 | NCDR | Classical T1D | f | [15-19] | 26.09 | Positive for GADA | 9.9 | 820 | Insulin | Type 2 diabetes in the family |
| 5 | NCDR | Likely T2D | f | [15-19] | 36.16 | Negative | 5.4 | 922 | OHA | N/A |
| 6 | NCDR | Classical T1D | m | [15-19] | 22.71 | Positive for GADA | 12.3 | 339 | Insulin | Type 2 diabetes in the family |
| 7 | NCDR | Classical T1D | m | [5-9] | 18.86 | Positive for GADA, IA-2A | 8.7 | 200 | Insulin | Sister with type 1 diabetes |
| 8 | NCDR | Classical T1D | m | [10-14] | 20.36 | Positive for GADA, IA-2A | 7,9 | 501 | Insulin | Mother with type 1 diabetes |
| 9 | NCDR | Classical T1D and MODY2 | m | [5-9] | 12.50 | Positive for GADA, IA-2A | 15.7 | 530 | Insulin | Brother, mother, maternal uncle and first cousin with hyperglycemia (MODY2) |

m: male; f: female; OHA: Oral hypoglycemic agents; N/A: not available in the registry
(*) Age at diagnosis: intervals to ensure the privacy and confidentiality of individuals: [0-4], [5-9], [10-14], [15,19], >19; (**) Auto- antibodies.

Supplementary Table 2. Clinical presentation of individuals with pathogenic deletions in $NCDR_{ALL}$ and $NMR_{ALL}$

| Gene | CNV | Registry | Subtype (prior analysis) | Sex | Age* (years) | BMI (kg/m2) | Aab** | HbA1c (%) | C-peptide (pmol/l) | Treatment | Family History of diabetes | Phenotype |
|---|---|---|---|---|---|---|---|---|---|---|---|---|
| *GATA4* | 8:8105359-11856903 | NMR | Known MODY | m | [10-14] | 18.33 | - | 10 | 86 | Insulin | N/A | Heart defect and congenital hernia |
| *HNF1A* | 12:120974510-121712218 | NMR | Likely MODY | m | > 19 | 24.48 | - | 11.1 | 524 | Insulin and OHA | Father, Paternal aunt and paternal grandfather have diabetes | N/A |
| | 12:121426776-121435265 | NMR | Likely MODY | f | [15-19] | 25.00 | - | 6.4 | N/A | Low dose of Insulin | N/A | N/A |
| | 12:121434074-121437802 | NMR | Known MODY | f | > 19 | 24.21 | - | 7.5 | N/A | insulin 10IUx2 | Mother, maternal grandmother, 3x maternal aunts and 1x maternal uncle, and a cousin | N/A |
| *HNF1B* | 17:34815551-36220373 | NMR | Known MODY | f | [0-4] | N/A | - | 5.3 | N/A | | N/A | Congenital kidney disease with bilateral kidney cysts and only one functioning kidney |
| | 17:34815551-36245768 | NMR | Known MODY | m | [15-19] | 28.02 | - | 8.5 | N/A | Insulin | No diabetes in the rest of family, however familiar hypomagnesemia | N/A |
| | 17:34815551-36245768 | NMR | Known MODY | f | > 19 | 23.59 | - | 5.6 | N/A | - | N/A | Hypomagnesemia, renal agenesis, Lynch Syndrome. Renal cysts., pancreatic calcifications, chronic diarrhoea |
| | 17:34815551-36249430 | NMR | Known MODY | m | [15-19] | 21.46 | - | 9.6 | N/A | Insulin | Mother, father and maternal grandmother had/have diabetes | N/A |
| | 17:34815551-36249430 | NMR | Known MODY | f | [15-19] | N/A | - | 7 | N/A | Insulin | Father with type 2 diabetes | Absent uterus. Mayer-Rokitansky-Küster-Hauser-syndrome. |

| | | | | | | | | | | | | |
|---|---|---|---|---|---|---|---|---|---|---|---|---|
| | 17:34815551-36249430 | NMR | Known MODY | m | [0.-4] | N/A | N/A | 4.3 | N/A | - | N/A | Renal cysts |
| | 17:34528650-36220373 | NCDR | Classical T1D | f | [5-9] | 16.24 | IA-2A | 7.4 | 480 | Insulin | N/A | N/A |
| | 17:34815551-36249430 | NCDR | Aab negative T1D | f | [15-19] | 21,33 | - | 6.5 | 840 | Insulin and metformin | No other individuals with diabetes in the family | N/A |
| | 17:34815551-36249430 | NCDR | Aab negative T1D | m | [15-19] | 18.78 | | 9.3 | 1041 | Insulin | N/A | Unspecified liver disease, allergy |
| | 17:34902695-36249430 | NCDR | Classical T1D | f | [15-19] | N/A | IA-2A | 11.9 | 392 | Insulin | N/A | N/A |

m: male; f: female; N/A: not available in the registry
(*) Age at diagnosis: intervals to ensure the privacy and confidentiality of individuals: [0-4], [5-9], [10-14], [15,19], >19.
(**) Autoantibody

Supplementary Table 3. Distribution of rare CNVs in $NMR_{CORE}$ and $NCDR_{CORE}$ after exclusion of pathogenic deletions and their carriers.

| Overlapped Regions | | CNVs MODY [516] | CNVs T1D [3270] | Frequency | | Association | |
|---|---|---|---|---|---|---|---|
| | | | | $NMR_{CORE}$ | $NCDR_{CORE}$ | OR (95%CI) | *P* |
| Global | DELs | 425 | 2602 | 0.82 | 0.80 | 1.20 (0.94, 1.53) | 0.14 |
| | DUPs | 371 | 2417 | 0.72 | 0.74 | 0.90 (0.73, 1.11) | 0.33 |
| Diabetes gene-set list | DELs | 0 | 5 | 0 | 0.02 | 0.63 (0.03, 11.60) | 0.76* |
| | DUPs | 2 | 10 | 0.004 | 0.003 | 1.27 (0.28, 5.81) | 0.76 |

(*) Pagano & Gauvreau 2X2 table correction for 0 values

Supplementary Table 4. Distribution of rare CNVs by interval size in $NMR_{CORE}$ and $NCDR_{CORE}$ after exclusion of pathogenic deletions and their carriers.

| | CNVs length | CNVs $NMR_{CORE}$ [516] | CNVs $NCDR_{CORE}$ [3270] | Frequency | | Association | |
|---|---|---|---|---|---|---|---|
| | | | | $NMR_{CORE}$ | $NCDR_{CORE}$ | OR (95%CI) | *P* |
| DELs | 50kb_100kb | 244 | 1383 | 0.47 | 0.42 | 1.22 (1.02, 1.47) | 0.03 |
| | 100kb_200kb | 118 | 844 | 0.23 | 0.26 | 0.85 (0.68, 1.06) | 0.15 |
| | 200kb_500kb | 53 | 300 | 0.10 | 0.09 | 1.13 (0.83, 1.54) | 0.43 |
| | 500kb_1000kb | 7 | 54 | 0.01 | 0.02 | 0.82 (0.37, 1.81) | 0.62 |
| | 1000kb_> | 3 | 21 | 0.01 | 0.01 | 0.90 (0.27, 3.04) | 0.87 |
| DUPs | 50kb_100kb | 133 | 1031 | 0.26 | 0.32 | 0.75 (0.61, 0.93) | 0.01 |
| | 100kb_200kb | 113 | 692 | 0.22 | 0.21 | 1.04 (0.83, 1.31) | 0.70 |
| | 200kb_500kb | 94 | 527 | 0.18 | 0.16 | 1.16 (0.91, 1.48) | 0.23 |
| | 500kb_1000kb | 23 | 126 | 0.04 | 0.04 | 1.16 (0.74, 1.83) | 0.51 |
| | 1000kb_> | 8 | 41 | 0.02 | 0.01 | 1.24 (0.58, 2.66) | 0.58 |

Supplementary Table 5. Rare duplications frequency distribution by interval size for individuals within the subtypes of classical T1D and autoantibody-negative T1D.

| CNVs length | CNVs Classical T1D [2954] | CNVs Aab_negative T1D [306] | Frequency | | Association | |
|---|---|---|---|---|---|---|
| | | | Classical T1D | Aab_negative T1D | OR (95%CI) | *P* |
| 50kb_100kb | 939 | 89 | 0.32 | 0.29 | 1.14 (0.88-1.47) | 0.33 |
| 100kb_200kb | 620 | 71 | 0.21 | 0.23 | 0.88 (0.66-1.16) | 0.37 |
| 200kb_500kb | 492 | 33 | 0.17 | 0.11 | 1.65 (1.14-2.40) | 0.007 |
| 500kb_1000kb | 111 | 14 | 0.04 | 0.05 | 0.81 (0.46-1.44) | 0.48 |
| 1000kb_> | 39 | 2 | 0.013 | 0.006 | 2.03 (0.49-8.46) | 0.32 |

Supplementary Table 6. Breakdown of the prevalence of different MODY subtypes based on genetic diagnosis in *known MODY* individuals

| MODY type | Known MODY + [107] | Known MODY +_- [194] | Known MODY + freq. | Known MODY +_- freq. | Two.prop test P | OR (95%CI) | P |
|---|---|---|---|---|---|---|---|
| INS-MODY | 2 | 3 | 0.02 | 0.02 | 1.00 | 1.21 (0.20-7.37) | 0.83 |
| MIDD-MT | 3 | 5 | 0.03 | 0.03 | 1.00 | 1.10 (0.26-4.65) | 0.91 |
| MODY1 - HNF4A | 4 | 11 | 0.04 | 0.06 | 0.65 | 0.64 (0.20-2.08) | 0.46 |
| MODY2 - GCK | 39 | 75 | 0.36 | 0.38 | 0.80 | 0.91 (0.56-1.48) | 0.71 |
| MODY3 - HNF1A | 49 | 85 | 0.46 | 0.44 | 0.83 | 1.08 (0.67-1.74) | 0.74 |
| MODY5 - HNF1B | 5 | 9 | 0.05 | 0.05 | 1.00 | 1.01 (0.33-3.08) | 0.99 |
| MODY8 - CEL | 3 | 3 | 0.03 | 0.02 | 0.75 | 1.84 (0.36-9.26) | 0.46 |
| MODY8 - CEL + T1D | 1 | 1 | 0.01 | 0.01 | 1.00 | 1.82 (0.11-29.41) | 0.67 |
| MODY - RFX6 | 1 | 1 | 0.01 | 0.01 | 1.00 | 1.82 (0.11-29.41) | 0.67 |
| MODY13 - KCNJ11 | 0 | 1 | 0.00 | 0.01 | 1.00 | 0.61 (0.24-15.00) | 0.76 |

+: rare deletions carriers

+_-: rare deletions carriers and non-carriers

INS-MODY: Insulin gene; MIDD-MT:Maternally Inherited Diabetes and Deafness maternally transmitted
